# A new system in qualitative RT-PCR detecting SARS-CoV-2 in biological samples: an Italian experience

**DOI:** 10.1101/2020.06.17.20124396

**Authors:** Marco Favaro, Walter Mattina, Enrico Salvatore Pistoia, Roberta Gaziano, Paolo Di Francesco, Simon Middleton, Silvia D’Angelo, Tullio Altarozzi, Carla Fontana

## Abstract

In the last moths the world was faced with the pandemic of a new severe acute respiratory syndrome coronavirus (SARS-CoV) and the majority of the Nations have yet to come out of it. Numerous assays have emerged to meet SARS-CoV-2 diagnostic needs. A clear knowledge of these assays’ parameters is essential to choose the proper test by clinical microbiologists. Unfortunately, the latter cannot be the unique criterion that guides test selection as - given the great demand - shortcomings of commercial kits is also a great issue. Aimed by the intention of overcoming both difficulties we have developed a new qualitative RT-PCR probe based for COVID-19 detection. The system detects three genes of SARS-CoV-2: RNA-dependent RNA polymerase (RdRp), envelope (E) and nucleocapsid (N) and β-actin gene used as endogenous internal control. The results of our assay show a total agreement with those obtained using a commercially available kit, with the exception of two specimens which did not pass the endogenous internal control. Moreover, our kit was designed to be open either for nucleic acid extraction step or on the RT-PCR assay to be carried out on several instruments. Thus, it is free from the industrial production logics of closed systems and conversely it is hypothetically available for distribution on large numbers in any microbiological laboratories. Presently, the kit is currently distributed worldwide

## INTRODUCTION

Since November 2019, the world has had to face an unprecedented public health emergency: the coronavirus: SARS CoV 2, now COVID-19 [1,2].

The pandemic has put a strain on the health system in its entirety and above all has put microbiologists in serious diagnostic difficulty, having to answer questions without having valid scientific evidence, especially in the early stages [3-5].

Over the time and through the gradual acquisition of new scientific findings - also supported by the World Health Organization (WHO) and Centers for Disease Control (CDC) and European Centers for Disease Control (ECDC) - evidences on the virus have become more and more available. The knowledge access has made possible to design diagnostic kits for the virus detection in biological samples. There have been many proposed diagnostic systems over the months that often differed in the proposed gene targets [6-9].

Gradually the various systems have passed the appropriate validations required by the Food and Drugs Administration (FDA) or CE IVD. However, the massive global spread of the virus has caused an objective difficulty in the supplying of diagnostic systems on the market, sometimes this is due to the viral nucleic acid extraction systems only, others to amplification and detection systems, frequently both [10].

The shortage of diagnostic systems has imposed a consequent limitation in their use, this has presented severe difficulties in having test kits available, which then resulted in delays in identifying positive patients. [10-12].

Unfortunately to date, even in our country, and with important differences from region to region, the problem persists.

On one hand, we have to deal with the population legitimate request to access the testing and their availabilities; on the other hand we have to face the same issue from health institutions and we do have to put up with the tech the technological and diagnostic resources actually available.

Furthermore, another key element is also the problem of limited human resources, engaged in microbiology laboratories to carry out the COVID19 tests.

After the moment of maximum spread of the virus, the real challenge for Italy as for the rest of the world, steered into the stocking of the diagnostic tests, the simplicity in the administration of the latter to the population and the sustainability of the system as a whole.

In a time of great diagnostic difficulty, our research team decided to design a new diagnostic system that could satisfy the required sensitivity and specificity, but allowing at the same time a reasonable rapid diagnosis, thus it could be introduced in a diagnostic process matching up to the requests of the health system. Our assay (from now kit) is a qualitative Reverse Transcriptase Real Time PCR probe-based (qRT-PCR) detecting target genes of COVID-19. In this paper we present the characteristics of our system.

## MATERIALS AND METHODS

### Specimens

A total of 166 nasopharyngeal swabs were included in the study. Samples were collected by trained staff using nasopharyngeal Eswab™ (Copan, Brescia-Italy) [13,14]. Once collected the swabs were quickly sent and processed at Life GeneMap Laboratory Messina (Italy) using a commercially available system: Novel Coronavirus (2019-nCoV) Real Time Multiplex RT-PCR Kit (Detection for 3 Genes) (LifeRiver, San Diego, CA 92121, USA). The leftover, remnants of specimens collected for routine clinical care or analysis that would otherwise have been discarded, were used in the evaluation of our system.

### Kit Design

Our diagnostic assay is a qualitative Reverse Transcriptase Real Time PCR probe-based (qRT-PCR). The targeted COVID-19 genes detected by our assay are: RNA-dependent RNA polymerase (RdRp), envelope (E) and nucleocapsid (N) of the COVID-19. Primers and Probes were designed on the base of the published sequence of COVID-19 on NCBI and were synthesized by Bio-Fab Research (Bio-Fab Research, Rome, Italy). Two set of primers, called “E” directed versus Envelop gene and “N” directed versus of Nucleocapsid gene, were specific to COVID19, one called RdpR, directed versus polymerase gene, was in common with SARS virus. The concentration of the primers and probes has been established by experimental procedure and the sensibility of the test performed with chimeric plasmid described below. Sequences of primers and Probes are not shown as patented.

A portion of an endogenous gene of human β-actin was used as internal control (IC) for the test, the latter also allowed the evaluation of correct nasopharyngeal sampling.

In order to evaluate our kit an initial proficiency assay was performed at Microbiology laboratories-Department of Experimental Medicine of “Tor Vergata” University of Rome. This proficiency-assay was run using a chimeric plasmids (CPs) in which sequences of the virus were artificially inserted on a plasmid [pBlueScript II SK(+)]. Synthesis of CPs was hired to Bio-Fab Research (Bio-Fab Research, Rome, Italy).

The specificity of the primers and probes was evaluated using ZeptoMetrix panels (ZeptoMetrix, Co.,Buffalo, NY,US): a)SARS-CoV-2 (E/ORF/1ab recombinant) Stock (ZeptoMetric), which is formulated with purified, intact bacterial cells containing synthetic SARS-CoV-2 sequences (the cells have been chemically modified to render them non–infectious and refrigerator stable); b) NATtrol™ Respiratory Verification panel 2 (ZeptoMetrix), which is formulated with purified intact virus particles and bacterial cells that have been chemically modified to render them non-infectious and refrigerator stable; c) NATtrol ™ Coronavirus-SARS Stock, containing intact virus particles modified to render them non–infectious and refrigerator stable. The panels are supplied in a purified protein matrix that mimics the composition of true clinical specimens.

The samples of ZeptoMetrix panels were treated as a common nasopharyngeal sample, but performed in triplicate. ZeptoMetrix tests were carried out at Adaltis Laboratories.

### Assay conditions

An aliquote 200μl of samples collected in Eswab™ was extracted both with manual procedure by silica magnetics beads procedure (MOLgen Universal Extraction Kit, QIAamp viral RNA) according to the manufacturer’s instruction. In order to process a large number of samples at time, the extraction procedure was also automatized on ExtraLab (Adalties srl, Guidonia, Italy).

The Real time amplification was performed in double on AmpliLab system (Adalties srl, Guidonia, Italy) and on CFX96 (Bio-Rad, Hercules, CA, USA) using qPCRBIO PROBE 1-Step Go No-Rox (PCR biosystems; www.pcrbio.com). In order to establish the proper amount of reverse transcriptase activity quantification of RTase Go was performed according to manufacturer’s instruction. Titration experiment showed that 0,2μl of 20X RTase Go, in amplification mix, gave good results in term of sensitivity.

Ideal condition of reaction was achieved using for 20 μl reaction a master mix composed as follow: 2X qPCRBIO probe 1-step Go Mix 10 ml; ppMix (Mix of all primer and probe) 5 μl; 20X Rtase Go 0,2 μl; and 5 μl of specimen. In particular ppMix contained as final concentration: 10 picomoles of RdpR, E and β-actin (Forward and Reverse primers), 30 picomoles of N (Forward and Reverse primer); 2,5 picomoles of probe for each target.

RT-PCR conditions, for both instruments, were the following: one step 45°C for 10 minutes; one step at 95°C for 2 min; 40 steps at 95°C for 5 seconds and the last one at 60°C for 25 seconds.

The instrument was programmed to read gene RdpR on Fam, gene E on Rox, gene N on Cy5 and in β-actin on Hex channel.

## RESULTS

The positivity in our test was based on WHO guidelines [6]. In particular, a sample was considered positive if showing a signal in at least one fluorophore Rox (gene E) and/or Cy5 (gene N), while the presence of a single positive signal on Fam channel was considered inconclusive, being the gene RdpR designed to be in common with other *Sabercoviridae*. On the contrary the absence of a signal on all channels, with the exclusion of Hex (that of β-actin), allowed us to conclude a sample as negative. The absence of a signal in all channels allowed us to conclude a sample as invalid due probable inhibition or an unreliable sampling. Table 1 reports the criteria of interpretation. Figure 1 reports the curve and the relative CT of a positive sample.

**Table 1.** Interpretation criteria used in our qRT-PCR assay

| Target gene and channel | Example 2 | Example 3 | Example 4 | Example 5 | Example 6 | Example 7 |
| --- | --- | --- | --- | --- | --- | --- |
| IC<br>(β-actin)<br>HEX | +/- | +/- | +/- | +/- | + | - |
| RdRPgene<br>(FAM) | + | + | + | + | - | - |
| E gene<br>(ROX) | - | + | + | - | - | - |
| N gene<br>(Cy 5) | - | + | - | + | - | - |
| Results interpretation | Negative for CoVid19<br>SabercoVirus detected | Positive for CoVid 19 | Positive for CoVid 19 | Positive for CoVid 19 | Negative | Sample invalid<br>probably inhibition or unsuitable withdrawal |

Note: Cut off value for all gene is $\leq 40$ ;
In presence of strong signal of others genes, the signal of IC may be inhibited but the results still valid.

**Figure 1.**
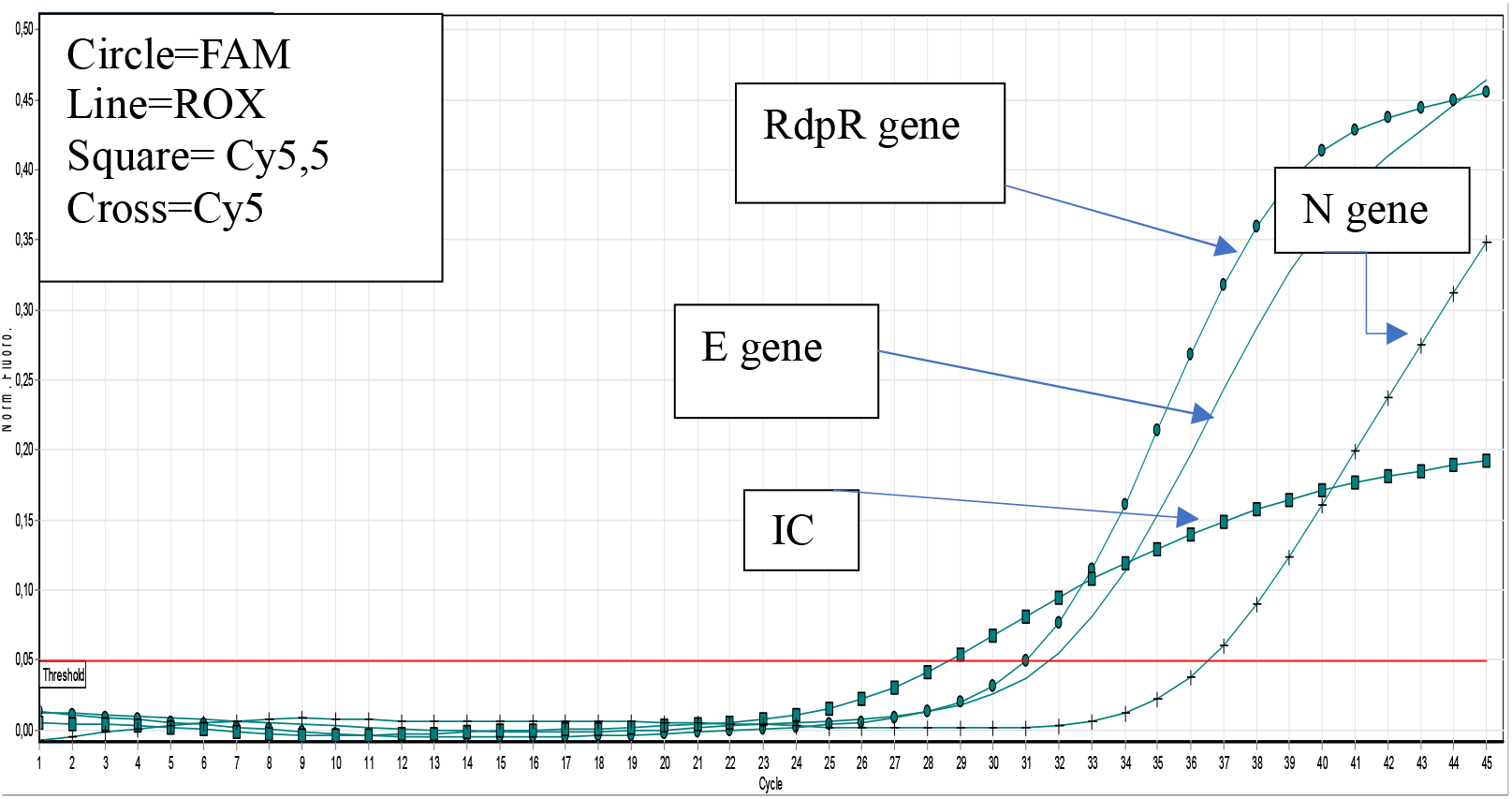
Image example of real-time RT-PCR curves of three target genes of SARC-CoV-2 and Internal Control (IC) detected by our assay.

The results of our test of specificity and cross reactivity tests, performed using the ZeptoMetrix panels, showed the absence of any cross-reaction as well as the specific reaction of our primers and probes toward COVID-19 genes (Figure 2).

**Figure 2.**
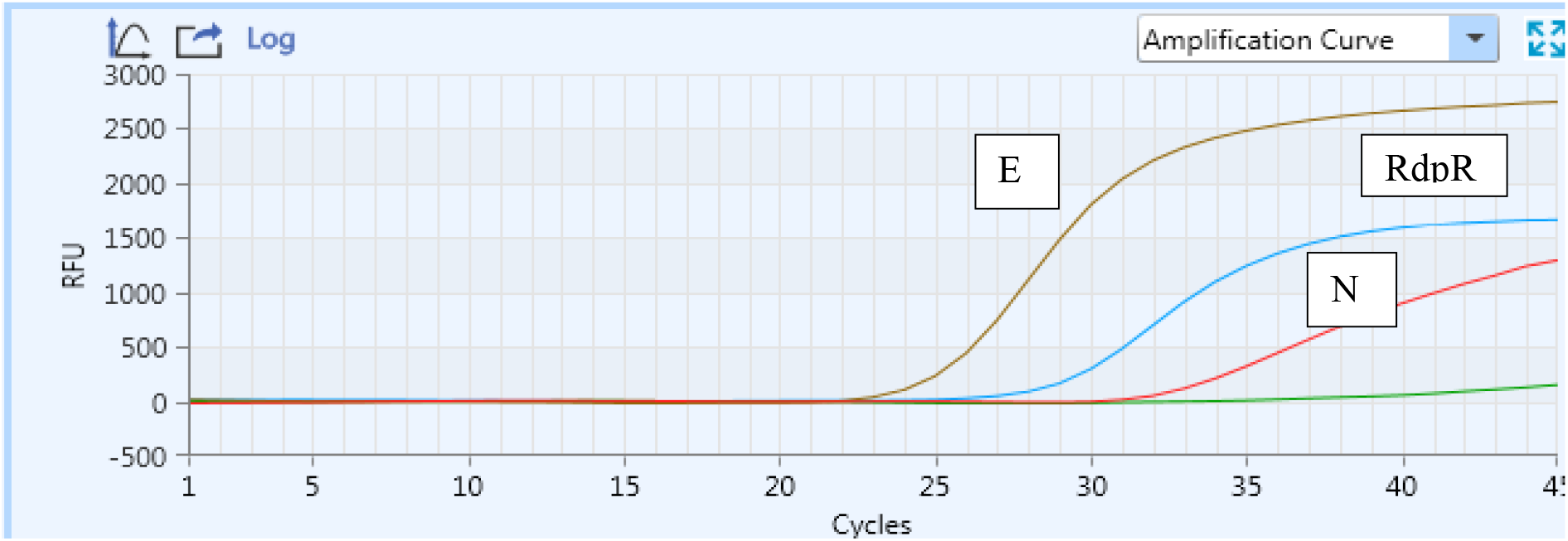
Results of qRT-PCR assays obtained using ZeptoMetrix ™ panels in detecting the target genes of SARS-CoV-2 virus.

In total 166 samples were included in the study. 133 samples were negative, 31 resulted positive, two samples did not show amplification of endogenous internal control (β−actin) and were concluded as indeterminate.

Among 31 positive sample, six were positive for RdRp only, therefore reported as “inconclusive”; and 25 were COVID-19 positive. Among these 25, nine were positive for all the targets while 16 RdpR and E only (see Table 2).

**Table 2.**
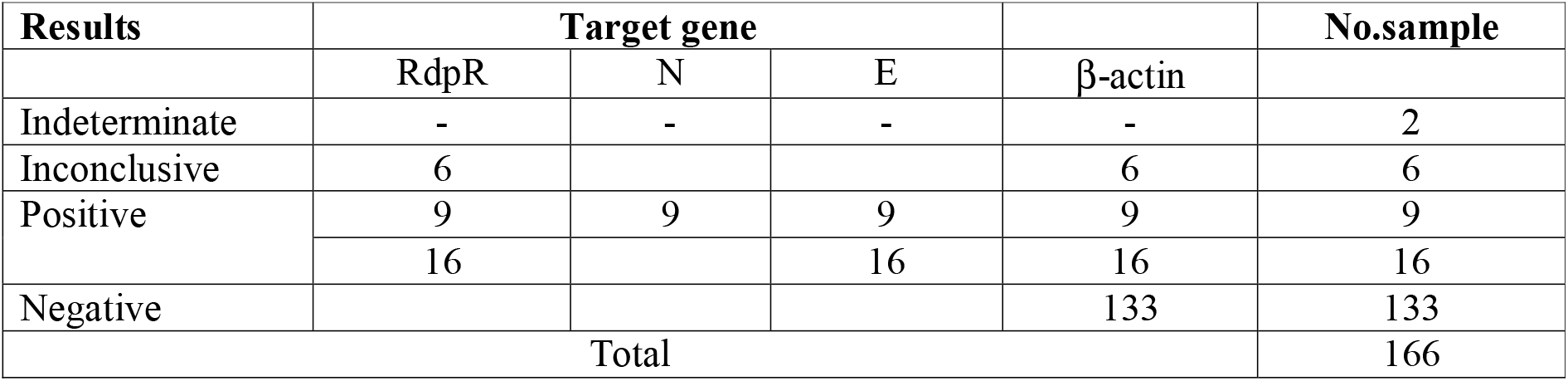
Results obtained by our assay (qRT-PCR).

| **Results** | **Target gene** |  |  |  | **No.sample** |
| --- | --- | --- | --- | --- | --- |
| | RdpR | N | E | β-actin | |
| Indeterminate | - | - | - | - | 2 |
| Inconclusive | 6 |  |  | 6 | 6 |
| Positive | 9 | 9 | 9 | 9 | 9 |
|  | 16 |  | 16 | 16 | 16 |
| Negative |  |  |  | 133 | 133 |
| Total |  |  |  |  | 166 |

The results showed by our assay were in agreement with those obtained using Novel Coronavirus (Novel-Cov19) Multiplex Real Time RT-PCR (RR-0479-02) (LifeRiver) for the initial diagnosis of SARS-CoV-2.

## DISCUSSION

Accurate and reliable diagnostic assays and large-scale testing are critical for early detection of a pathogen involved in an outbreak, but it is more crucial in case of an epidemic event for prompt public health actions. This has proven to be particularly true for SARS-CoV-2, which was identified as the cause of an outbreak of pneumonia, in Wuhan (China) in December 2019, and quickly spread all over in the world [15-21]. Laboratory diagnosis of infections caused by severe acute respiratory syndrome coronavirus 2 (SARS CoV-2) is mainly accomplished by performing nucleic acid amplification tests (NAATs) on respiratory tract specimens. Indeed, the upper respiratory tract specimens such as nasopharyngeal swabs and oropharyngeal swabs generally have high SARS-CoV-2 viral loads upon symptom onset [22]. Some authors have recently suggested to extend NAAT testing in order to include saliva and stool samples [23,24]. Due to the pressing worldwide request of tests to diagnose COVID-19, numerous SARS-CoV-2 NAAT assays have become available and many others are in the final stages of development [20, 28, 29]. This aspect has the great advantage of making a wide range of diagnostic tests available to health systems and therefore allows us to respond to the diagnostic needs caused by the pandemic, but on the other hand this incredible quantity and difference between each kit poses problems on the validation process itself. An important restriction to validation qRT-PCR molecular assays for detection of SARS-CoV-2 is the availability of virus RNA standard and debate is still ongoing as whether or not to consider integral SARS-CoV-2 RNA full length as a safety level 3 biological hazard [19,20]. If debate reaches consensus on marking SARS-CoV-2 as Level 3, its treatments could only be performed by laboratories with appropriate level 3 (BSL3) security measures, whose number is scarce thus limiting the capacity of carrying out experimental tests [28]. Taking into account these critical issues we have developed a qRT-PCR assay able to detect three target genes of SARS CoV-2 which strongest aspect is the characteristics of IC which is able to evaluate jointly the accurate collection of nasopharyngeal samples and the presence of any inhibitors in the PCR reaction. Sampling in SARS-CoV-2 is, in fact, one of the most frequent and probable cause of false negative results and therefore of delayed diagnosis [15,28]. Moreover, in order to avoid to work with a full-lenght viral RNA, overcoming the issue of BSL3, we have chosen to construct artificial chimeric plasmid to test our primes and probes and at the same time to use ZeptoMetrix panels which allowed us to evaluate the specificity of our assay in safe conditions.

Our kit was designed to be open, either for nucleic acid extraction step or on the RT-PCR assay to be carried out on several instruments (in the present paper we tested two of them). Therefore, our assay can be used in any molecular biology laboratory. Furthermore, our kit is free from the industrial production logics of “closed systems” and conversely it is hypothetically available for distribution on large numbers. This aspect, at a time of great demand for tests and of equally known shortcomings of commercial kits, can be a significant strength that facilitates the introduction into microbiological laboratories.

In addition, taking into account potential genetic drift of SARS-CoV-2, especially as the virus evolves within new populations, and albeit literature suggests at least two molecular specific targets should be included in the assay to reduce the likely of cross-reactions, we are implementing our kit by adding a forth target gene codifying for glycoprotein spike (S) S gene, but validation tests are still in progress [25-29]. Finally, as evidences emerged in literature show that aside to direct respiratory sampling, rectal swab as well as saliva could be proper specimens to enhance the diagnosis of COVID-19, we are also extending validation test of our assay on such direction [28].

## Data Availability

the data used in the manuscript are avaivailable if required

## Acknowledgments

This study beneficed of a financial support by Adaltis s.r.l. The funders had no role in study design, data collection and interpretation, or the decision to submit the work for publication.

## Ethics statement

Ethical approval was not required, having based this study on the use of leftover human specimens collected for routine clinical care or analysis that would otherwise have been discarded. The same specimens are “unlinked anonymized materials”. This statement in agreement to FDA “Guidance on Informed Consent for In Vitro Diagnostic Device Studies Using Leftover Human Specimens that are Not Individually Identifiable” April 25, 2006 and “Bioetica ed uso dei campioni biologici umani” Pezzati P. & Graziani MS biochimica clinica, 2008, vol. 32, n. 3.

## Conflict of Interest

Carla Fontana has received a research grant by Quintiles/Angelini. Advisory Board: Angelini, Pfizer

Marco Favaro has received a research grant by Alifax R&D and Adaltis s.r.l

Walter Mattina, Simon Middleton, Silvia D’Angelo and Tullio Altarozzi are employed in Adaltis

